# Multivariate Analysis of the Impact of Demographic and Clinical Factors on Cardiovascular Health

**DOI:** 10.1101/2024.05.19.24307595

**Authors:** Youssef Benfallah, Wanru Guo, Shoaib Sarfraz, Yanyu Yang

## Abstract

**Background:** This study investigates the influence of demographic and clinical factors on cardiovascular health using a multivariate analysis approach. We analyzed data from one of the largest heart-related disease datasets, merging variables such as age, sex, chest pain type, to determine their association with resting blood pressure, cholesterol, and maximum heart rate.

**Methods:** Two-means comparison is used to investigate the effect of heart disease with response variables, One-way and two-way MANOVA are used to study the relationship between chest pain type, as well as sex and heart disease on the response variables, respectively. Profile analysis and MANCOVA were also conducted to determine a final model.

**Results:** Our findings indicate significant effects of heart disease on resting blood pressure, cholesterol and maximum heart rate. Chest pain type also has a significant effect on maximum heart rate achieved, but not blood pressure and cholesterol. There are also notable interactions between sex and heart disease status on all response variables. Finally, we formulized a multivariate regression model with age, sex, chest pain type, heart disease, and the interaction between heart disease and sex as predictors.

**Conclusion:** These findings can inform physicians on possible treatment strategies in cardiovascular diseases.

## 1 Introduction

Cardiovascular diseases (CVDs) remain the leading cause of mortality worldwide, claiming approximately 17.9 million lives annually (American College of Cardiology, 2023). This staggering statistic underscores the urgent need for enhanced predictive strategies to identify individuals at high risk of developing these conditions. This study aims to leverage a comprehensive dataset sourced from multiple international databases, to understand the influence of demographic variables such as age, sex, and specific clinical presentations like chest pain type on cardiovascular outcomes.

The complexity of cardiovascular disease mechanisms suggests that a multifaceted analysis approach is required to fully understand the interplay between these variables and cardiovascular health outcomes. The integration of multiple heart-related databases provides a unique opportunity to examine these relationships across a diverse patient population, thus potentially unveiling patterns that are not observable in more homogenous datasets (Yamada et al, 1997).

Furthermore, multivariate analysis offers a robust framework for examining the simultaneous effects of multiple independent variables on several dependent variables (Wilhelmsen et al, 1973). This statistical approach is ideal for exploring the effects of demographic and clinical factors on outcomes such as resting blood pressure, cholesterol levels, and maximum heart rate— critical indicators in the management and prognosis of heart disease. Such comprehensive analyses are pivotal, not only for understanding the immediate impacts of these variables but also for their long-term consequences on patient health (Provenzano et al., 2023).

This analysis will explore these relationships, with the goal of informing clinical practice and possibly guiding future research in cardiovascular health management. By understanding how various factors interact to influence cardiovascular risk, this study hopes to contribute to more tailored and effective interventions, thus reducing the global burden of these deadly diseases.

## 2 Methods

### Study Design/Data Source

The study is a retrospective analysis design utilizing an aggregated dataset compiled from five leading heart-related disease databases: Cleveland (OH), Hungary, Switzerland, Long Beach (VA), and Statlog. The final dataset comprised 918 unique patient records after removing duplicates. Predictors of interest were Age (years), Sex (Male, Female), Chest Pain Type (TA, ATA, NAP, ASY), and Heart Disease (0 = none, 1 = present). Outcomes of interest were RestingBP (mmHg), Cholesterol (mm/dL), and MaxHR (bpm). Full variable descriptions are listed in Table A1. Missing values for resting blood pressure and cholesterol, coded as a value of 0, were removed from the dataset. Histograms of continuous variables, as well as box plots of outcomes by categorical groups can be found in Figures A1-A9. Additionally, scatterplots of outcome variables by age are shown in Figures A10-A12.

### Models

All statistical analyses were conducted using R. The statistical tests used for this analysis include comparison of two means, one-way multivariate analysis of variance (Eq AO1), two-way MANOVA, profile analysis, and multivariate covariance analysis (MANCOVA). Due to its versatility, a large sample Hotelling’s T^2^ test was used to compare mean vectors between those with and without heart disease. A one-way MANOVA was performed to analyze differences in outcomes between each of the four chest pain types (Eq AO3). A two-way MANOVA was performed to check for possible interactions between heart disease status and sex. A profile analysis was conducted to identify possible parallel, coincident, or level relationships for heart disease status and sex. Lastly, a MANCOVA model was used to identify relationships for the full set of variables in this study. The mathematical forms of each model are listed in the Appendix. All tests were performed at the α = 0.05 significance level.

### Assumptions

Assumptions for the statistical tests included independence, multivariate normality, and equal covariance matrices. The independence assumption was satisfied by the original study design, where each observation represents a unique participant. Chi-square plots were generated for each level of the categorical variables, including cell-wise interactions between heart disease status and sex. Combined with the Chi-square plots, multivariate Shapiro-Wilk’s tests were performed to check the normality assumption. Box’s M-test was used to check for equal covariance matrices between groups.

Chi-square plots did not show significant deviation from the reference line for heart disease, sex, and three chest pain types (ASY, NAP, ASY). The Chi-sq plots indicated non-normality for the TA chest pain type group and all four interactions between heart disease and sex. The multivariate Shapiro-Wilk’s tests indicated non-normality for both heart disease groups (no: p < 0.0001, yes: p < 0.0001), both sex groups (F: p < 0.0001, M: p < 0.0001), three of the four chest pain types (TA: p < 0.0001, ASY: p < 0.0001, NAP: p < 0.0001, ATA: p = 0.5388), and three of the four interaction groups (F w/o HD: p < 0.0001, F w/ HD: p = 0.0627, M w/o HD: p < 0.0001, M w/ HD: p < 0.0001). Due to the large sample size of the data set, we can still assume normality.

Box’s M-test indicated violation of the equal covariance assumption for sex (p = 0.0418), chest pain type (p = 0.0248), and cell-wise heart disease and sex interaction (p = 0.0394). Heart disease (p = 0.0565) met the equal covariance assumption. Given the importance of the equal variance assumption and its sensitivity to non-normality, a log transformation and square root transformation was attempted, but did not show any improvement in multivariate normality.

## 3 Results

### 3.1 Comparison of Two Means

The large sample Hotelling’s T^2^ test was performed with resting blood pressure, cholesterol, and maximum heart rate to identify significant differences between individuals with and without heart disease.

Post-hoc analysis was performed using simultaneous and Bonferroni-corrected confidence intervals generated for each outcome.

### 3.2 One-way MANOVA

Chi-square plots of each chest pain type are generated (**image AO5)** All plots exhibit a decent fit at lower quantiles but show various degrees of divergence at higher quantiles. The asymptotic chest pain type plot shows a great in the mid to high quantile range. These observations could be symptomatic of non-normality in the distributions. A Multivariate Shapiro-wilk test was conducted (**Table AO6)**. The rejection of normality in three (ATA, NAP, ASY) out of the four groups suggests the need for large sample approach testing, in addition to careful interpretation of the equal variance assumption results. The equality of covariance matrices across each chest pain type group was assessed using a Box’s M test (**Table 3**).

**Table 1.** Hotelling’s T^2^ test, Large Sample Approach.

| Test | $T^2$ | crit value | df | p-value |
| --- | --- | --- | --- | --- |
| Two-Mean Vector Comparison | 147.4911 | 7.8147 | 3 | $< 0.001^*$ |

**Table 2.** Simultaneous and Bonferroni-corrected Confidence Intervals.

| $\bar{x}_{i1} - \bar{x}_{i2}$ | Simultaneous | Bonferroni |
| --- | --- | --- |
| Resting BP | (-9.4977, -2.4831) * | (-8.994, -2.9868) * |
| Cholesterol | (-24.4224, -0.1627) * | (-22.6802, -1.9049) * |
| Max HR | (13.8588, 23.158) * | (14.5266, 22.4902) * |

**Table 3.** One Way MANOVA Equal Variances Assumption Test.

| Test | M | $\chi^2$<br>Critical value | df | P-value |
| --- | --- | --- | --- | --- |
| Box's M | 32.00679 | 31.563 | 18 | 0.02475 |

**Table 4.** Large Sample One-Way MANOVA Test Result.

| Test | Wilks' ($\Lambda^*$) | $\chi^2$<br>Score | $\chi^2$<br>Critical value | Df | $\alpha$ | p-value |
| --- | --- | --- | --- | --- | --- | --- |
| One-Way<br>MANOVA | 0.84 | 127.51 | 16.92 | 9 | 0.05 | < 0.001 |

The results from Box’s M test indicate that the null hypothesis of equal covariance matrices can be rejected (p = 0.02475). Given the importance of the equal variance assumption and its sensitivity to non-normality, a log transformation and square root transformation was also attempted but did not show any improvement in multivariate normality.

The One-Way MANOVA was conducted using the large sample approximation method (E**q AO3**), to explore the impact of chest pain type on resting blood pressure, cholesterol levels, and maximum heart rate.

The null hypothesis was rejected, indicating a significant chest pain type effect on the responses (E**q AO4**). A Post hoc ANOVAs test was conducted to further investigate the differences among the groups for each dependent variable (**Table AO7**). All three responses reject the null hypothesis, with an extremely low p-value for the “MaxHr” response.

Given the possible relationship between the responses and the relatively higher p-values for RestingBp and Cholesterol, Bonferroni corrected Confidence intervals are generated the adjusted alpha level 0.0056, based on an initial alpha of 0.01 (**Table AO4**).

The Bonferroni corrected CI demonstrate that only intervals for the response “MaxHr” do not contain zero. Subsequent ranking of groups was performed in **table 5** for the response “MaxHr”.

**Table 5.**
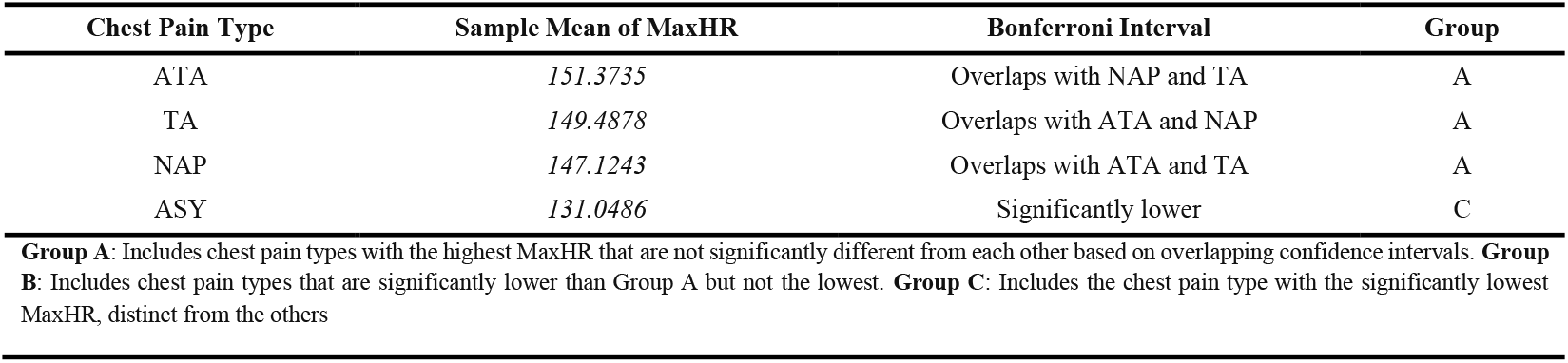
Final Grouping Table.

**Table 6.**
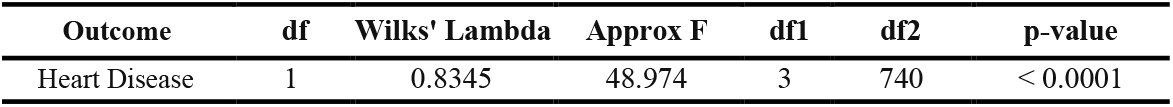
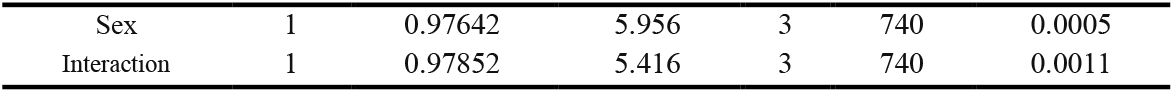
Two-way MANOVA.

| Outcome | df | Wilks' Lambda | Approx F | df1 | df2 | p-value |
| --- | --- | --- | --- | --- | --- | --- |
| Heart Disease | 1 | 0.8345 | 48.974 | 3 | 740 | < 0.0001 |
| Sex | 1 | 0.97642 | 5.956 | 3 | 740 | 0.0005 |
| Interaction | 1 | 0.97852 | 5.416 | 3 | 740 | 0.0011 |

**Table 7.**
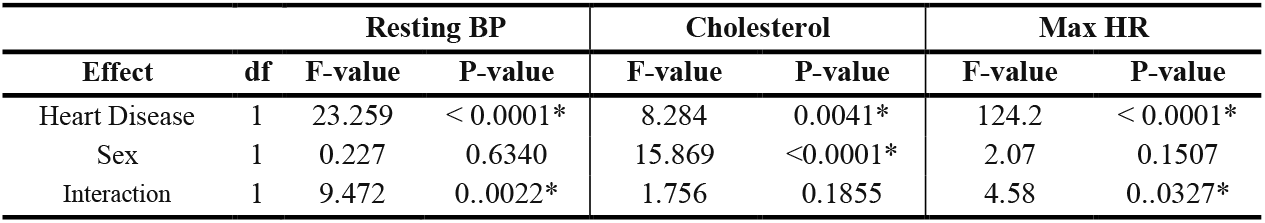
Post-hoc analysis – Univariate two-way ANOVA.

**Table 8.** Test for the Significance of Age.

| Test | $\chi^2$ Score | crit value | df | p-value |
| --- | --- | --- | --- | --- |
| FM vs RM | 106.0773 | 7.81 | 1 | < 0.001 |

**Table 9.** Parallel tests for both profiles.

| Test | Test Statistic | Critical Value |
| --- | --- | --- |
| Heart Disease | 140.3405 | 6.0237 |
| Sex | 22.64 | 6.0237 |

Group A, consisting of ATA, NAP, and TA, includes chest pain types with the highest MaxHR, showing no significant differences among them. Group C, represented solely by ASY, exhibits a significantly lower MaxHR compared to all other types, distinctly setting it apart.

No Group B is identified in this scenario as no chest pain types fall between the high MaxHR of Group A and the significantly lower MaxHR of Group C.

**Figure 1.**
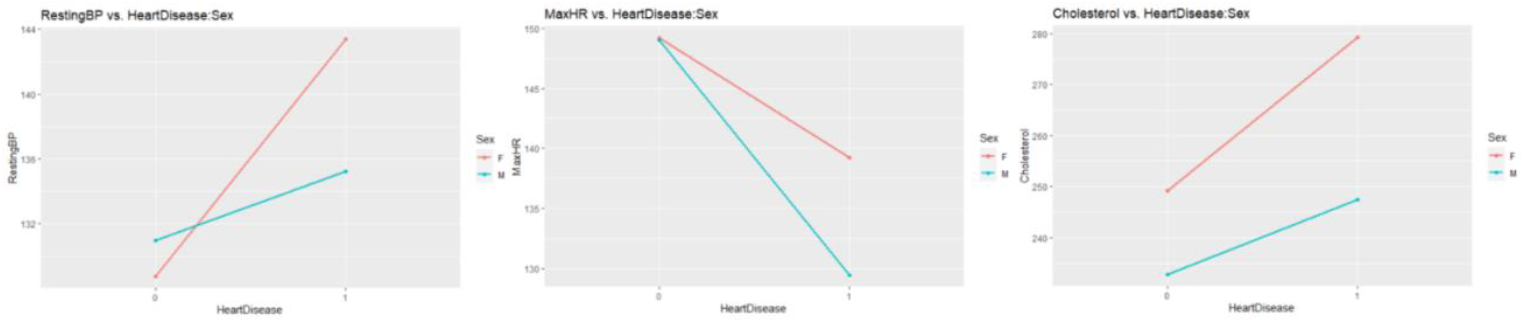
Interaction effects of heart disease and sex on outcomes.

### 3.3 Two-way MANOVA

The two-way MANOVA indicates a significant interaction between heart disease status and sex. On average, males with heart disease had lower resting blood pressure, maximum heart rate, and cholesterol than females with heart disease. Males without heart disease had slightly higher resting blood pressure, similar maximum heart rate, and lower cholesterol than females without heart disease. Both males and females with heart disease had higher resting blood pressure, cholesterol, and lower maximum heart rate than males and females without heart disease. However, since the interaction is significant, we conducted post-hoc analyses to interpret the main effects on each response variable.

The post-hoc analysis reveal a significant interaction effect between heart disease and sex on resting blood pressure and maximum heart rate, but not cholesterol. Heart disease and sex individually had significant effects on cholesterol. Specifically, having heart disease and being female significantly increases cholesterol levels (interaction plot).

### 3.4 MANCOVA

A test with nested models was conducted to test if the variable age significantly improves the model containing sex, chest pain type, heart disease, and the interaction between heart disease and sex.

The test statistic (106.0773) is much larger than the critical value (7.814728) for one degree of freedom at the usual 0.05 significance level. Therefore, the null hypothesis is rejected. Age significantly improves the model.

**Figure 2.**
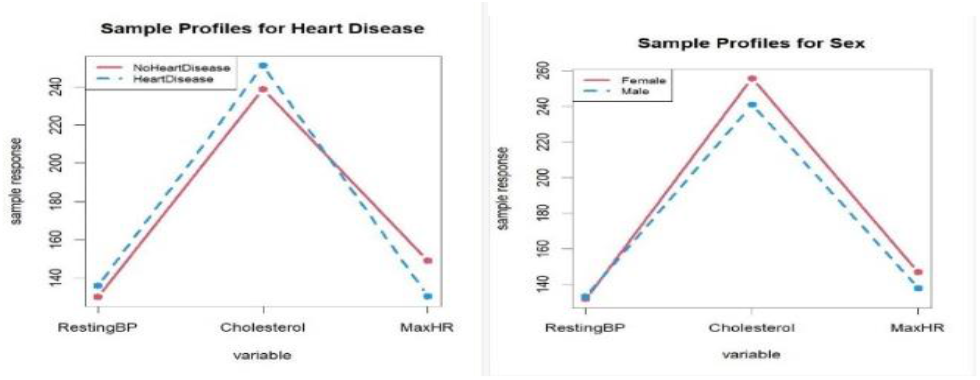
Sample profiles for both heart disease and sex.

### 3.5 Profile Analysis

Since both test statistics were higher than the critical value at alpha value 0.05, we reject the null for both. We did not continue the profile analysis to coincident or level profiles, and conclude that none of the profiles have a parallel pattern.

## 4 Discussion

The results of the Two Mean Comparison Test showed that there is a statistically significant difference in all of RestingBP, Cholesterol, and MaxHR between people with or without Heart Disease, with the Post-Hoc Analysis’ confirming it. It showed that those without heart disease demonstrated significantly lower resting blood pressure and cholesterol than those with heart disease. Conversely, those without heart disease had a higher maximum heart rate than those with heart disease.

The application of one-way MANOVA in our study clearly demonstrated that chest pain type significantly influences the responses. The result of Wilks’ Lambda (Λ = 0.84) with a highly significant chi-square score (χ^2^ = 127.51) and p-value (< 0.001) comfirms it. The subsequent one-way ANOVA post-hoc analyses further delved into the influence of chest pain type on individual cardiovascular indicators. Although the analysis revealed variations in resting blood pressure and cholesterol levels, they were not statistically significant after Bonferroni correction. In contrast, maximum heart rate showed significant differences, highlighted by an F-value of 40.73 and a p-value of <0.0001, suggesting this parameter’s acute sensitivity to the type of chest pain experienced. Bonferroni corrected confidence intervals refined these findings. Indeed, MaxHR differences were statistically significant between ASY (Asymptomatic) and the other types (ATA and NAP), This finding indicates that asymptomatic individuals may not exhibit the expected cardiovascular response under stress, potentially underestimating their cardiovascular risk. The strategic grouping based on the MaxHR differences, showed in Group A (Atypical Angina, Non-Anginal Pain, Typical Angina), where no significant differences were found among these chest pain types regarding MaxHR, a comparable pathophysiological stress response, which could imply similar underlying mechanisms or treatment approaches. Lastly, Resting Blood Pressure and Cholesterol did not show significant differences across chest pain types after correction, suggesting that these cardiovascular indicators might be influenced by additional factors not solely explained by chest pain type.

The Two-Way MANOVA results highlight the complex interplay between heart disease, gender, and the cardiovascular health indicators (RestingBP, Cholesterol,MaxHR). Despite significant interaction effects between gender and heart disease in the multivariate model making it difficult to interpret the main effects, we had some interesting findings in the post-hoc analysis with each univariate variable. The significant interaction effects observed for RestingBP and MaxHR underscore the necessity of considering both heart disease status and gender when evaluating cardiovascular risks or planning treatments. Specifically, the analysis suggests that men and women may experience different cardiovascular responses when affected by heart disease, which could have implications for personalized medical strategies. As evident in the interaction plots, females with heart disease are likely to have higher resting BP and cholesterol levels, posing increased cardiovascular risks compared to their male counterparts. The MaxHR findings are particularly concerning as show that heart disease significantly lowers maximum heart rate, a critical marker of cardiovascular health. This effect was more pronounced in males. Despite the significant findings for RestingBP and MaxHR, the lack of interaction effect on Cholesterol suggests that while having heart disease and being female individually increases cholesterol levels, their combined impact does not differ significantly from their independent effects.

Additionally, the inclusion of age in the MANCOVA model decisively enhances the model’s explanatory power. Indeed, test statistic of 106.0773 significantly exceeds the critical value of 7.81 (p < 0.001), The finding confirms age as a crucial variable in understanding and managing cardiovascular health effectively. This underlines the need for age-specific considerations in medical research. Finally, in the profile analysis we did not find any parallel pattern between either different heart disease or sex groups, which confirms both our two-mean comparison and two-way MANOVA tests that those with heart disease having a lower cholesterol level, blood pressure and higher max heart rate than those without heart disease, and females having higher cholesterol and maximum heart rate than males.

We do recognize a few limitations for this study. Firstly, there are multiple variables in the dataset, however we only considered age, sex, chest pain type and heart disease. Other variables such as ECG findings and blood sugar level all play important roles in cardiovascular parameters. We could have used backwards model selection/AIC to determine the best model starting from a model with all parameters (including all interactions), which would likely result in a lower AIC and better predictive performance than our current model. Secondly, our data were unbalanced between different groups and many of our normality assumptions were violated, which calls for the need of cautious interpretation of our findings. The study’s reliance on large sample assumptions due to deviations from normality, and unequal covariances from the two-mean comparison test also means that further studies would be needed to confirm our findings. Lastly, our dataset was created by merging 5 datasets from different geographical locations, which raises concerns for the generalizability of our findings. Further study would be needed to confine our cohort to a specific population to achieve more reliable findings.

## 5 Conclusion

Based on all the statistical tests performed, a consistent result that has emerged is that Age, Sex, Chest Pain Type, and Heart Disease are all significant predictors of cardiovascular outcomes. Our main findings are that there are significant effects of heart disease on resting blood pressure, cholesterol, and maximum heart rate. Chest pain type also has a significant effect on maximum heart rate, but not resting blood pressure and cholesterol. Additionally, there are significant interactions between sex and heart disease status on all the response variables of this study. All taken together, these findings can guide clinicians in personalized risk assessment, treatment planning, and preventative strategies. It will help clinicians know what demographic or medical factors to prioritize over others when looking for the most important people to target with their preventative campaigns.

## Data Availability

https://www.kaggle.com/datasets/fedesoriano/heart-failure-prediction

