## Appendix for "Multivariate Analysis of the Impact of Demographic and Clinical Factors on Cardiovascular Health"

### Appendix (Benfallah *et al.* 2024)

**Table A1. Overview of Study Variables and Their Descriptions**

| Variable Name | Type | Description | Role in Study |
|---|---|---|---|
| Age | Continuous | Age of the patient in years | Independent Variable |
| Sex | Categorical | Sex of the patient (Male, Female) | Independent Variable |
| Chest Pain Type | Categorical | Chest pain type (TA, ATA, NAP, ASY) | Independent Variable |
| Heart Disease | Binary | Presence of heart disease (1 = present, 0 = none) | Independent Variable |
| RestingBP | Continuous | Resting blood pressure (mm Hg) | Dependent Variable |
| Cholesterol | Continuous | Serum cholesterol (mm/dl) | Dependent Variable |
| MaxHR | Continuous | Maximum heart rate achieved | Dependent Variable |

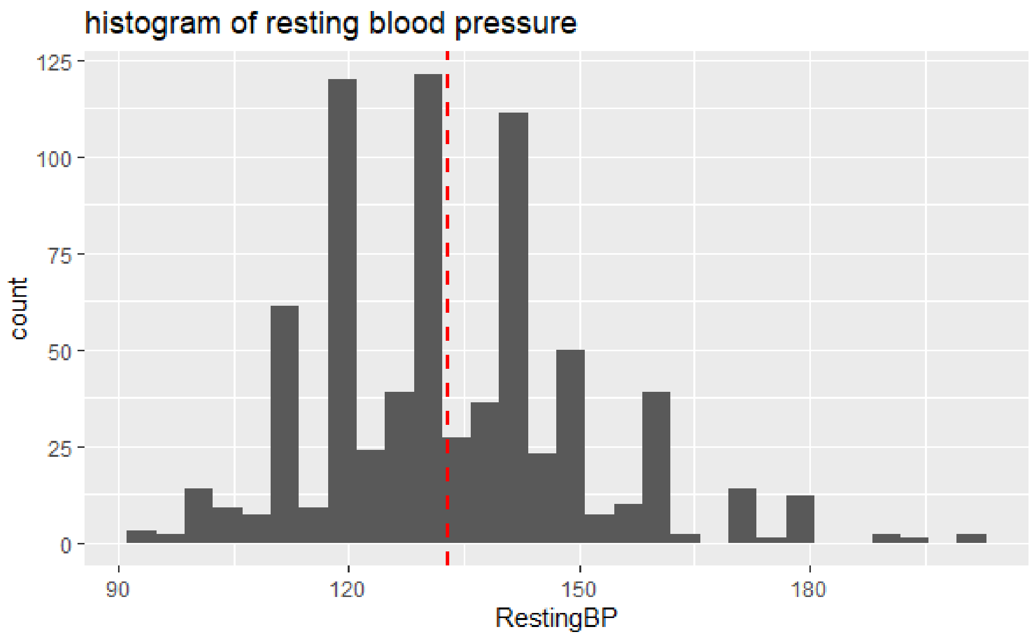


**Figure A1.** Histogram of resting blood pressure.

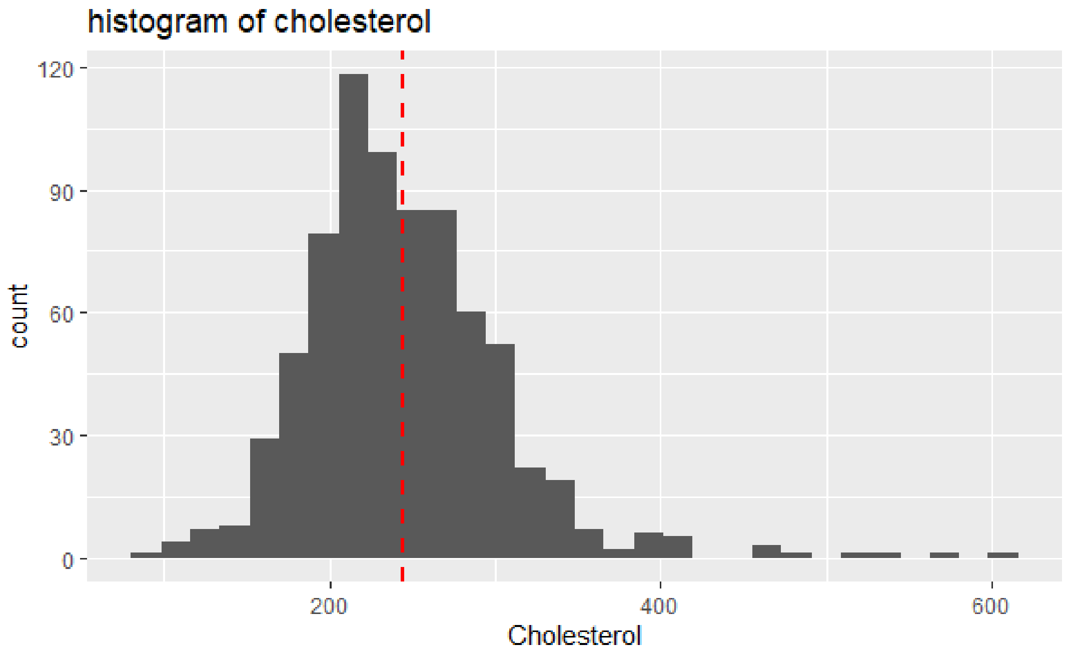


**Figure A2.** Histogram of cholesterol.

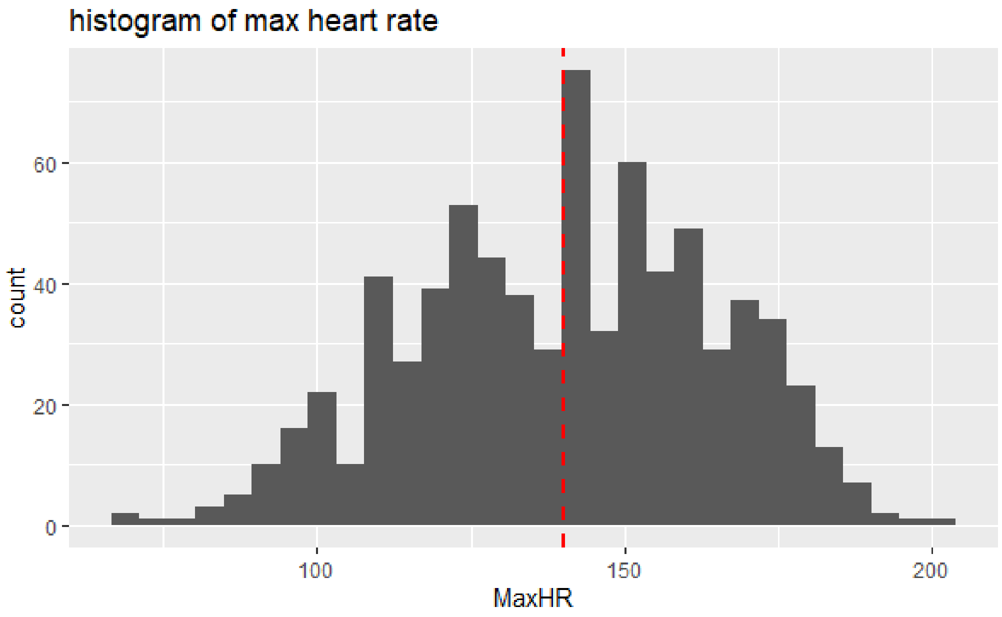


**Figure A3.** Histogram of maximum heart rate.

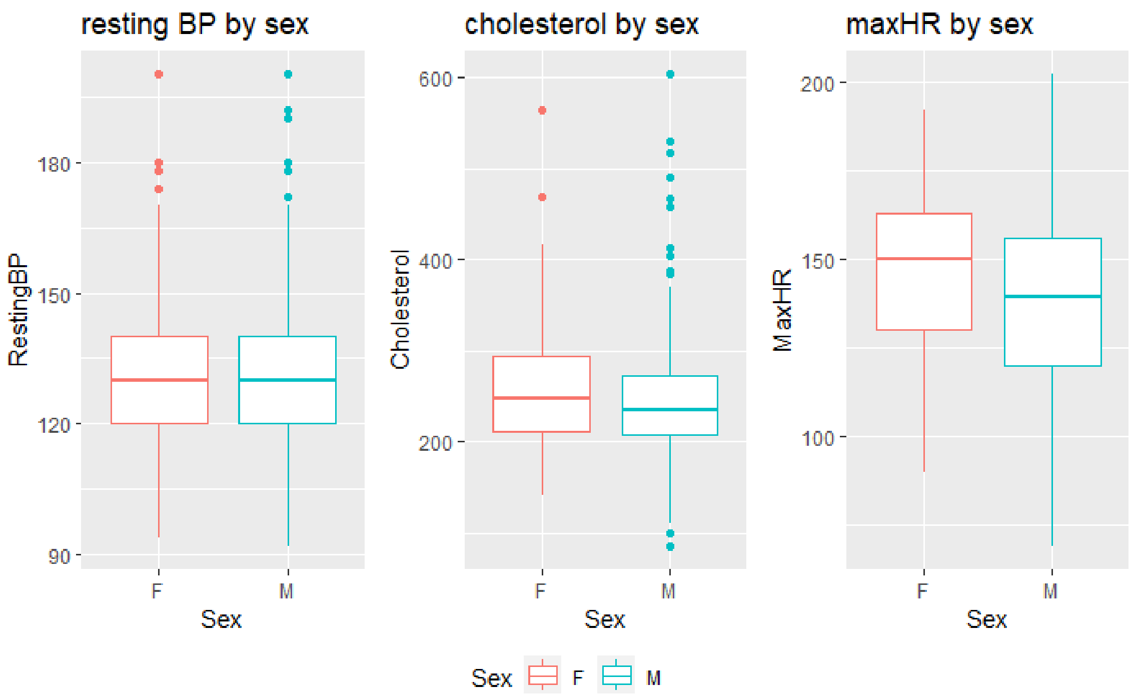


**Figure A4.** Box plots of outcome variables by sex.

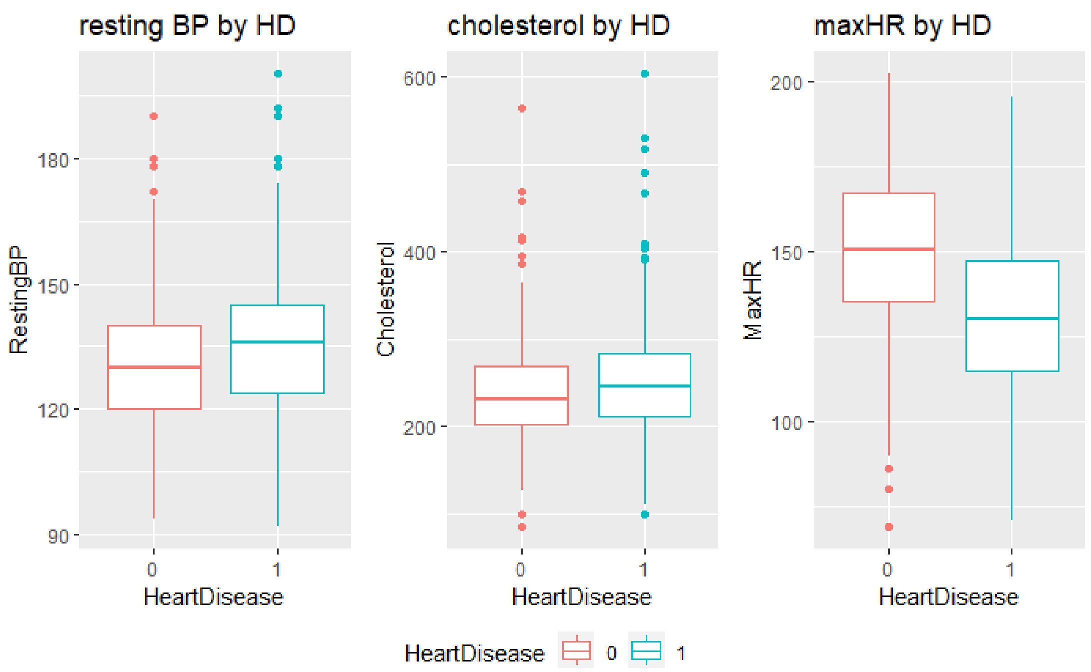


**Figure A5.** Box plots of outcome variables by heart disease status.

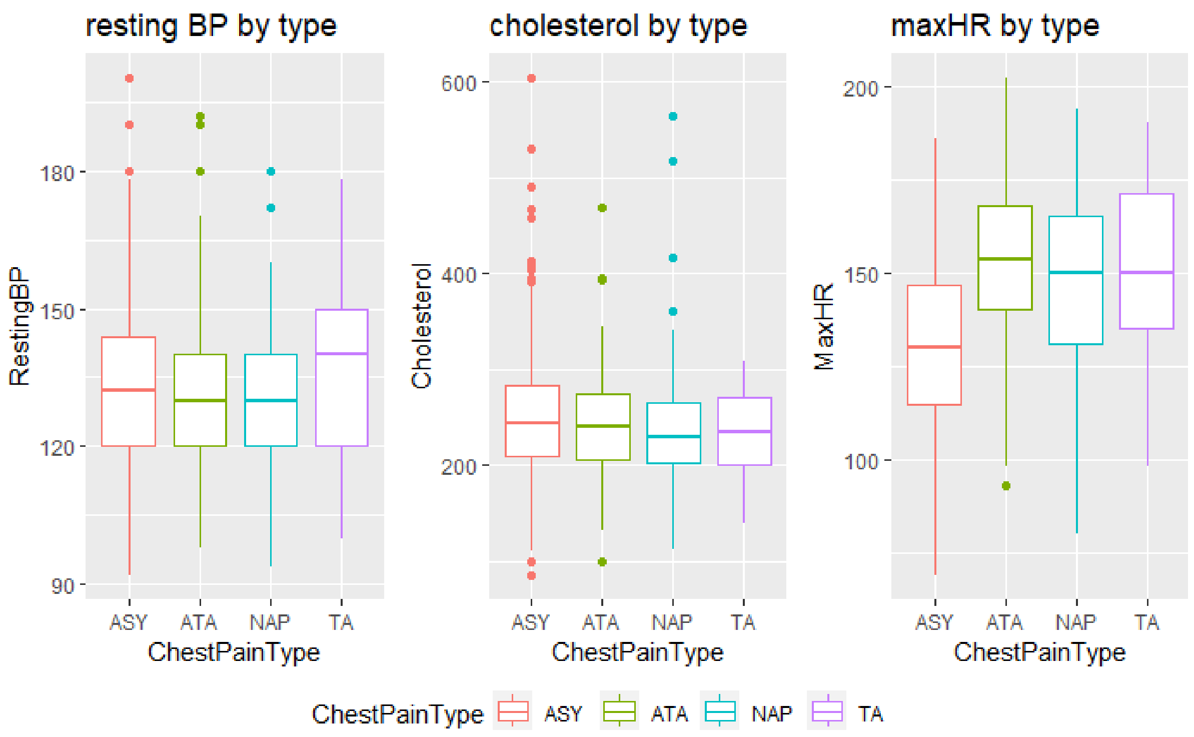


**Figure A6.** Box plots of outcome variables by chest pain type.

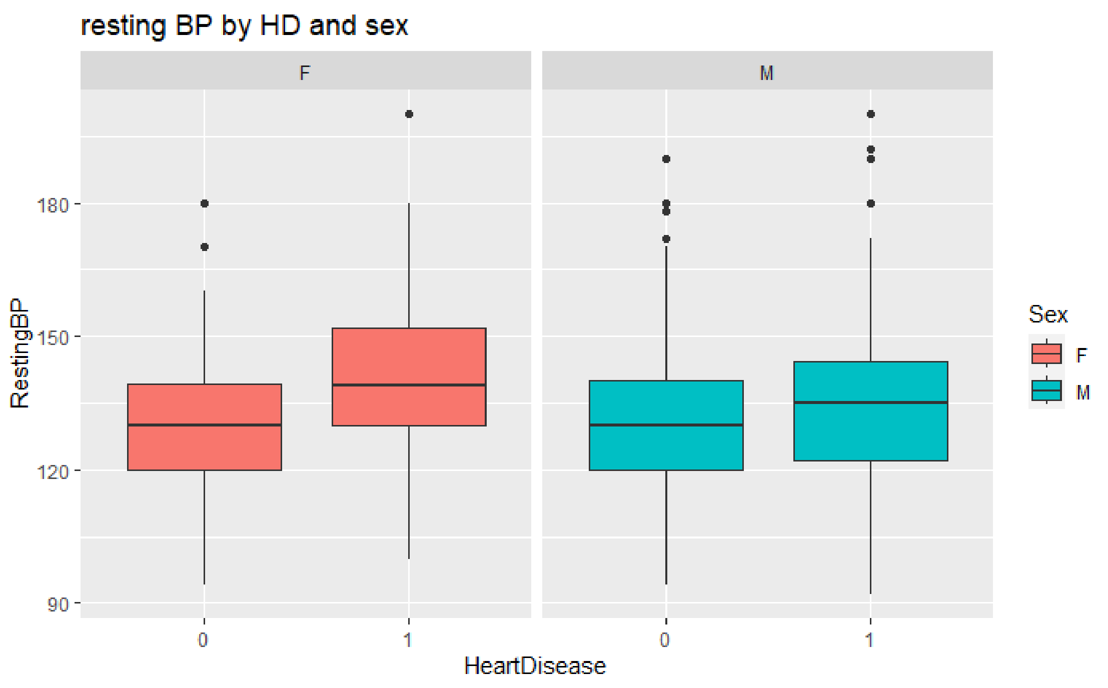


**Figure A7.** Resting blood pressure by heart disease status and sex.

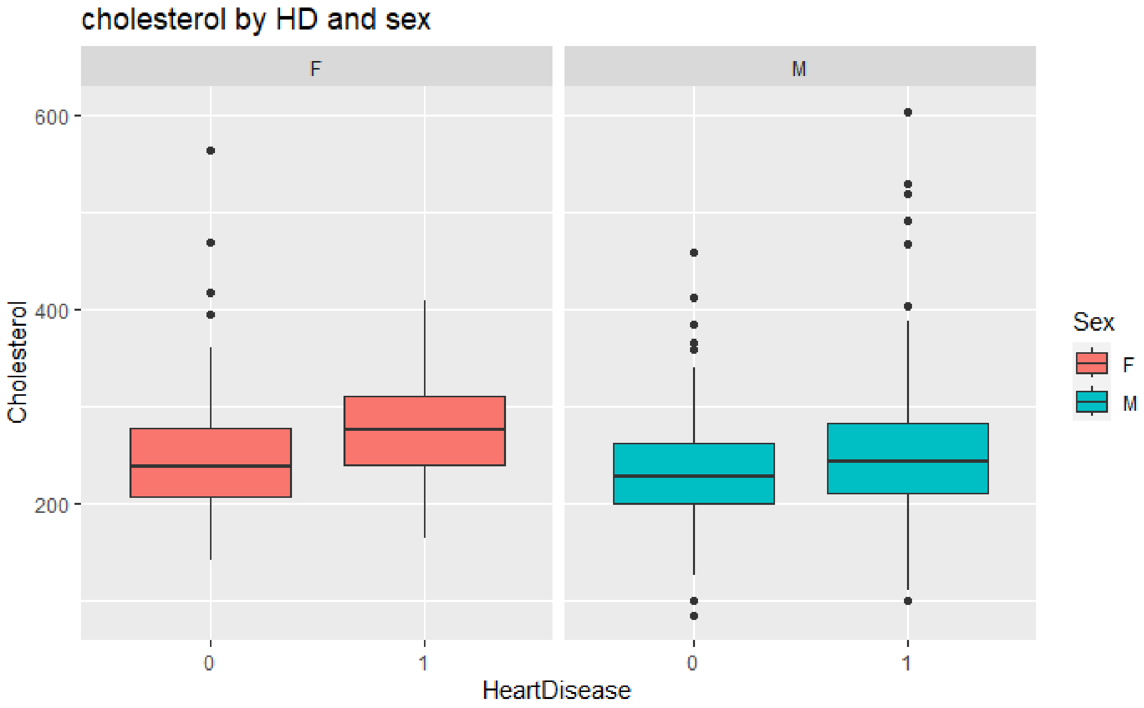


**Figure A8.** Cholesterol by heart disease status and sex.

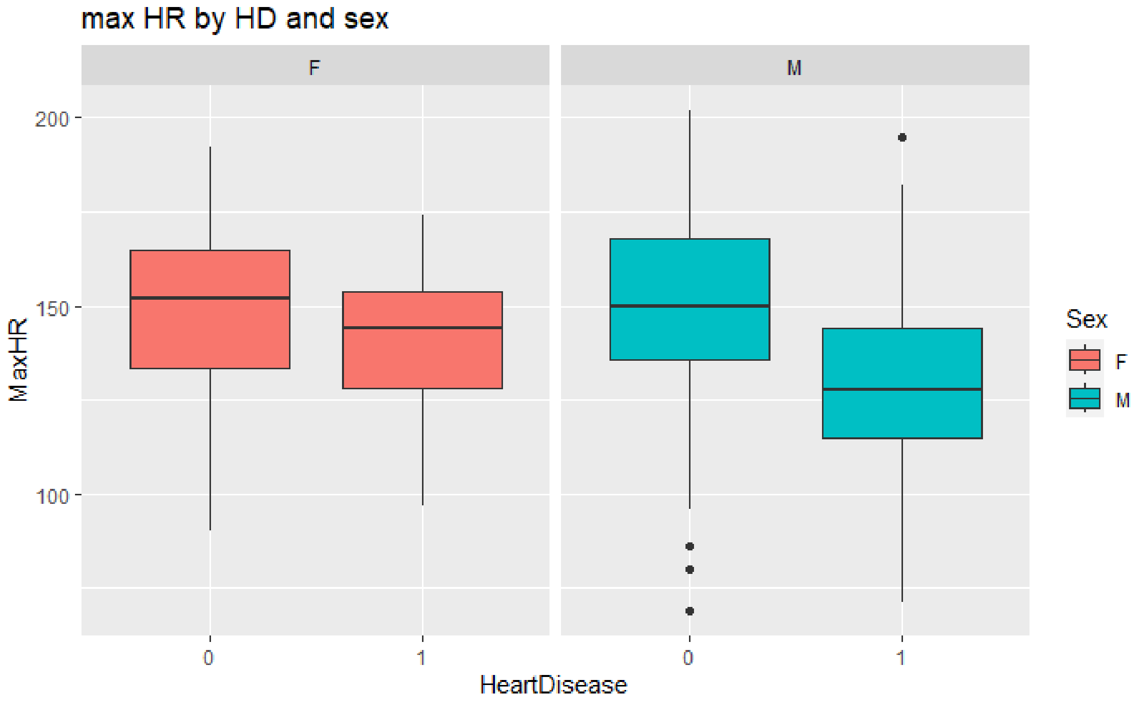


**Figure A9.** Maximum heart rate by heart disease status and sex.

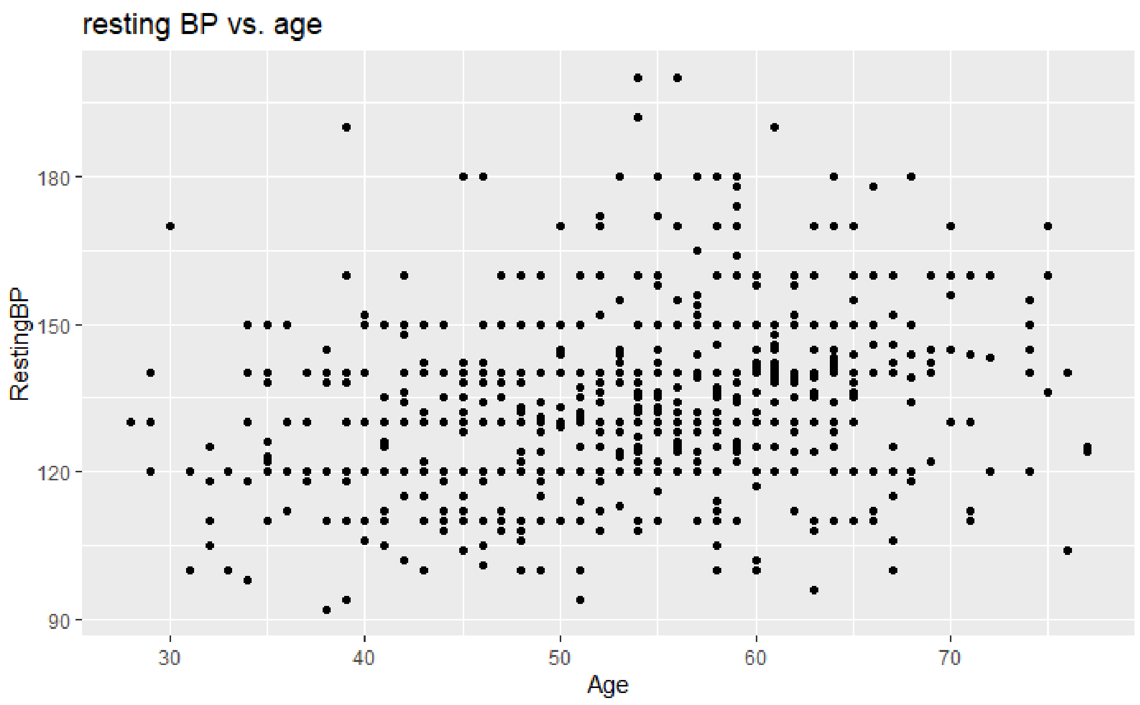


**Figure A10.** Scatter plot of resting blood pressure by age.

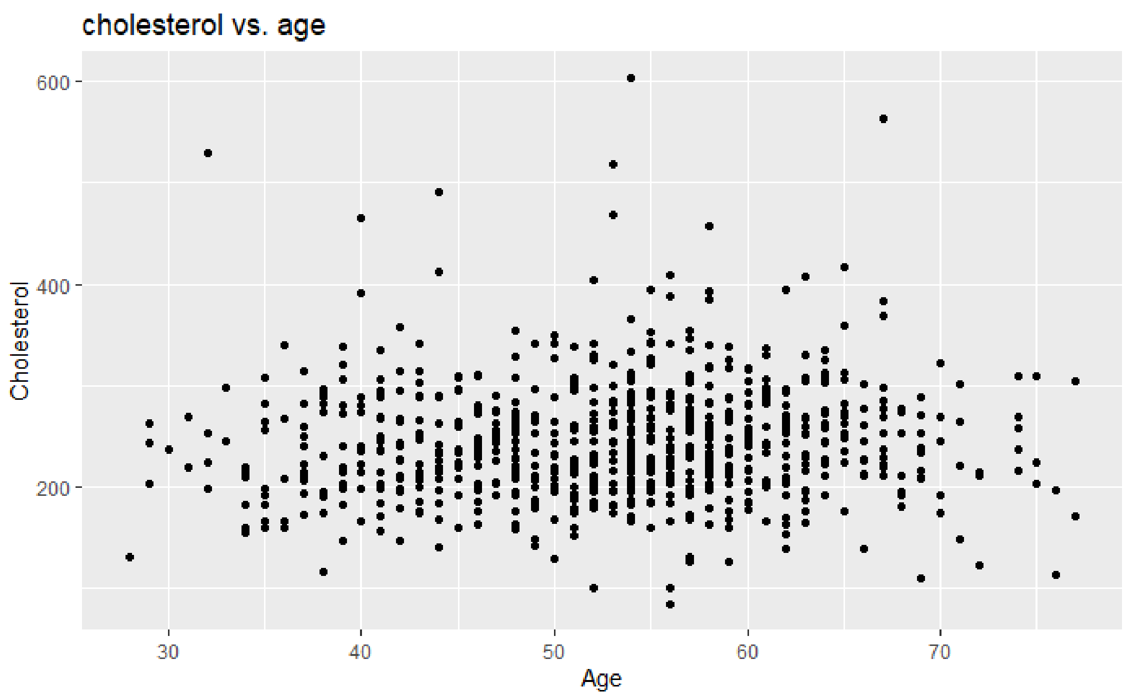


**Figure A11.** Scatter plot of cholesterol by age.

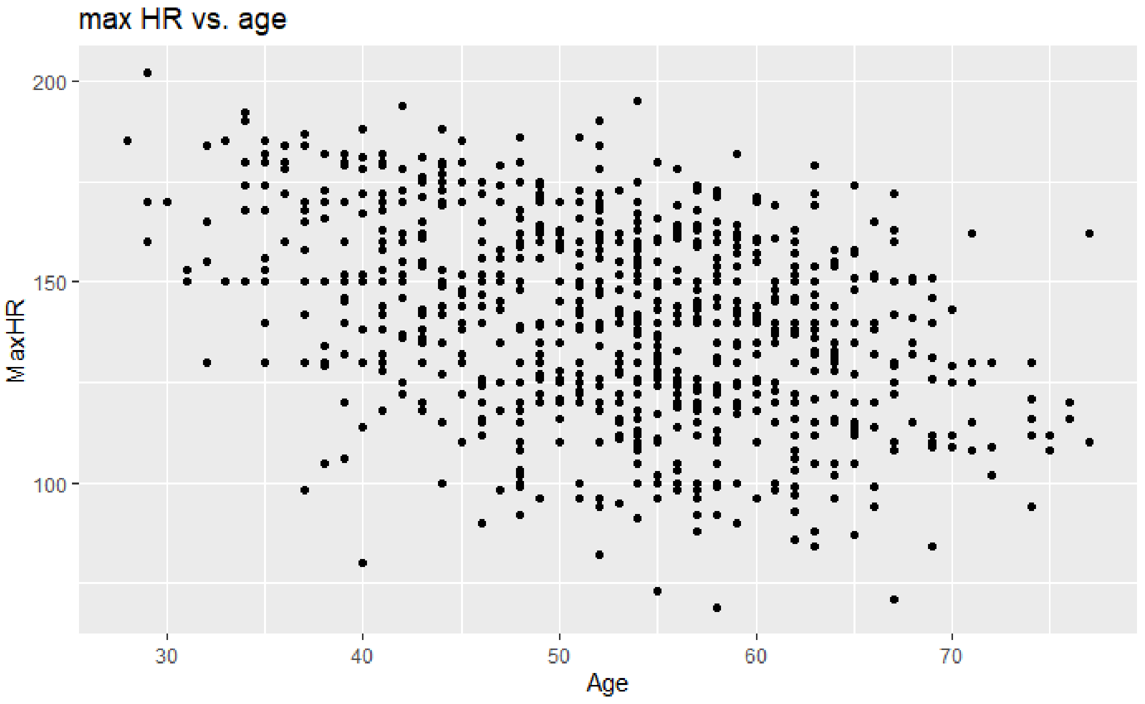


**Figure A12.** Scatter plot of maximum heart rate by age.

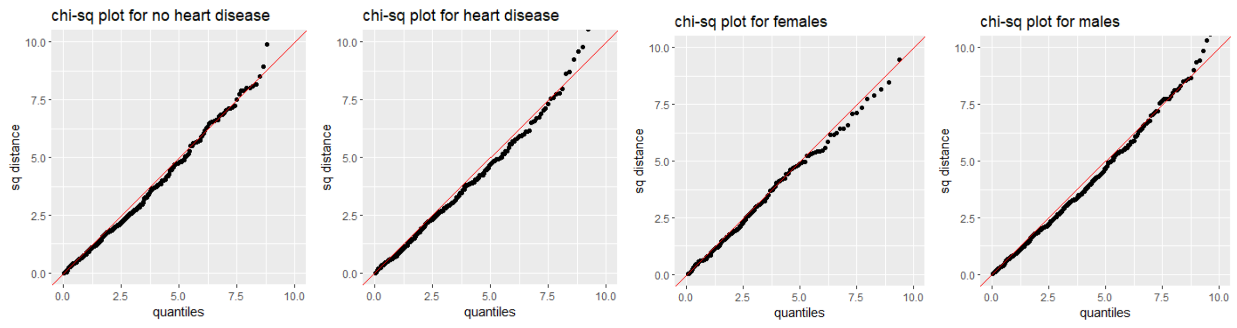


**Figure A13.** Chi-square plots for heart disease and sex. (L-R: no HD, HD, females, males)

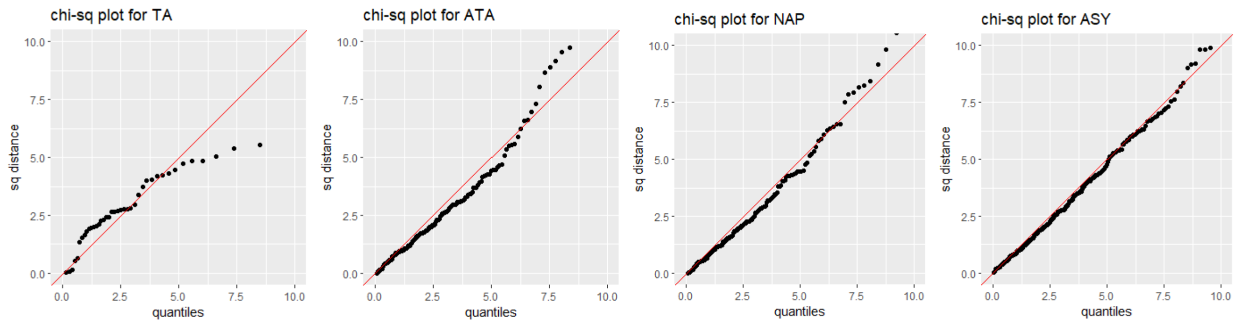


**Figure A14.** Chi-square plots for chest pain type (L-R: TA, ATA, NAP, ASY)

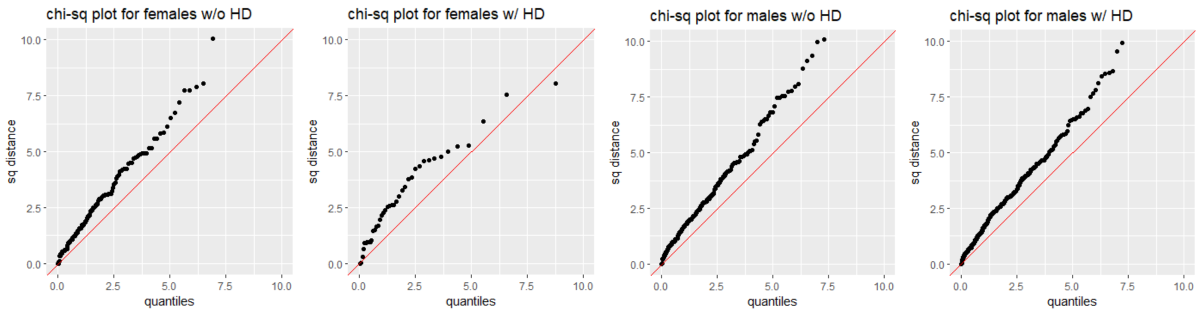


**Figure A15**. Chi-square plots for heart disease and sex interaction. (L-R: F w/o HD, F w/ HD, M w/o HD, M w/HD)

$$\left(T^2 = \left(\overline{\boldsymbol{X}}_1 - \overline{\boldsymbol{X}}_2\right)^T \left(\frac{\boldsymbol{S}_1^2}{n_1} + \frac{\boldsymbol{S}_2^2}{n_2}\right)^{-1} \left(\overline{\boldsymbol{X}}_1 - \overline{\boldsymbol{X}}_2\right)\right)$$

**Formula 1.** Formula for Welch’s t test, used for testing differences between groups with and without heart disease

$$Y_{ij} = \mu + \tau_i + \boldsymbol{e_{ij}}$$

Where $\boldsymbol{Y_{ij}}$ is the vector of observed measurements for resting BP, cholesterol, and MaxHR for subject $j$ in chest pain type, $\boldsymbol{\mu}$ is the overall mean vector for all dependent variables. $\tau_i$ is the effect vector of the $i$ -th chest pain type on the dependent variables. $e_{ij}$ is the vector of random errors associated with subject $j$ in chest pain type $i$ ..

**Model 1.** Model for the One Way MANOVA approach

$$\Lambda^* = \frac{|W|}{|B+W|}$$

**Formula 2.** Formula for Wilks' Lambda, derived from the determinant of matrices representing the sum of squares and cross-products (SSCP) of both within-group variations (|W|) and the total variation (|B+W|).

$$-\left(n - 1 - \frac{p+g}{2}\right)\ln(\Lambda^*) \sim \chi^2_{p(g-1)}$$

**Formula 3.** Formula for large sample version of the test is performed to assess whether the groups significantly differ in their multivariate means, the test statistic is computed by transforming (Λ*) using a logarithmic function, which approximates a chi-square distribution.

$$X_{ijk} = \mu + \tau_i + \beta_j + (\tau\beta)_{ij} + e_{ijk}$$

where:

$X_{ijk}$ represents the dependent variables (resting BP, cholesterol, MaxHR) for each observation.
$\mu$ is the overall mean.
( $\tau_i$ ) represents the effect of the $i$ heart disease type.
($\beta_j$) represents the effect of the $j$th gender.
$(\tau\beta)_{ij}$ represents the interaction effect between the $i$th heart disease type and the $j$th gender.
($e_{ijk}$) is the random error term.

**Model 2.** Model for the Two Way MANOVA approach

[illegible]

$$\Lambda^* = \frac{|SSCP_{residuals}|}{|SSCP_{effect} + SSCP_{residuals}|}$$

**Formula 4.** Formula for The Likelihood Ratio Test (LRT). This is used to determine the significance of the heart disease, sex and interaction effects. This test uses the ratio of the determinants of the sum of squares and cross-product matrices (SSCP), provides a statistic

$$-\left(gb(n-1) - \frac{p+1-df_{effect}}{2}\right)\ln(\Lambda^*) \sim \chi^2_{df_{effect} \cdot p}$$

**Formula 5.** Formula for The logarithm of the ratio obatined from Formula 4 is then used to form a test statistic that approximates a chi-square distribution.

The first model is:

- $X_{lj} = [X_{lj1}, X_{lj2}, X_{lj3}]'$, where:
  - variable indicator i=1,2,3 (RestingBP, Cholesterol, MaxHR, respectively)
  - Group indicator, l=1,2 (Heart Disease or No Heart Disease respectively)
  - Observation indicator, j=1,2,3.......,$n_l$ (n1=390, n2=356)

The second model is:

- $X_{lj} = [X_{lj1}, X_{lj2}, X_{lj3}]'$, where:
  - variable indicator i=1,2,3 (RestingBP, Cholesterol, MaxHR, respectively)
  - Group indicator, l=1,2 (Females or Males respectively)
  - Observation indicator, j=1,2,3.......,$n_l$ (n1=182, n2=564)

**Model 3.** Model for Profile Analysis. The first model is for Heart Disease, the second is for Sex.

$$(\overline{x_1} - \overline{x_2}) \cdot C' \cdot \left[\left(\frac{1}{n1} + \frac{1}{n2}\right) \cdot C \cdot Spooled \cdot C'\right]^{-1} \cdot C(\overline{x_1} - \overline{x_2})$$

**Formula 6.** Formula is for calculating the test statistic for Parallel tests of Profile Analysis'

$$\frac{((n1+n2-2) \cdot (p-1))}{n1+n2-p} \cdot F_{p-1, n1+n2-p}(\alpha)$$

**Formula 7.** Formula is for calculating the critical value for Parallel tests of Profile Analysis'

**AO (One Way MANOVA Annex):**

**AO1:**

we consider a vector of responses $Y_{ij}$ for each subject $j$ in chest pain type $i$. The model is formulated as follows:

$$Y_{ij} = \mu + \tau_i + \boldsymbol{e_{ij}}$$

Where $\boldsymbol{Y_{ij}}$ is the vector of observed measurements for resting BP, cholesterol, and MaxHR for subject $j$ in chest pain type, $\mu$ is the overall mean vector for all dependent variables. $\tau_i$ is the effect vector of the $i$ -th chest pain type on the dependent variables. $e_{ij}$ is the vector of random errors associated with subject $j$ in chest pain type $i$ ..

## AO2:

$$\Lambda^* = \frac{|W|}{|B+W|}$$

Determinant of matrices representing the sum of squares and cross-products (SSCP) of both within-group variations (|W|) and the total variation (|B+W|).

## AO3:

To ensure robustness against non-normality, a large sample version of the test is performed to assess whether the groups significantly differ in their multivariate means, the test statistic is computed by transforming (Λ*) using a logarithmic function, which approximates a chi-square distribution.

$$-\left(n - 1 - \frac{p+g}{2}\right) \ln(\Lambda^*) \sim \chi^2_{p(g-1)}$$

Where "n" is the count of all individuals assessed across the various categories of chest pain. "p" is the count of responses (RestingBP, Cholesterol, and MaxHR). "g" is the count of types of chest pain. If the calculated chi-square from exceeds the critical value at the desired level of significance (0.05), The null hypothesis is rejected.

## A04:

**Table AO4.** Bonferroni-corrected confidence intervals by chest pain type

| Response Variable | Comparison | Confidence Interval | Statistically Significant? |
|---|---|---|---|
| **Cholesterol** | ATA vs NAP | *-12.20371 to 23.63117* | No |
| | ATA vs ASY | *-22.64908 to 7.98527* | No |
| | ATA vs TA | *-18.03859 to 39.15143* | No |
| | NAP vs ASY | *-28.26867 to 2.17739* | No |
| | NAP vs TA | *-23.70200 to 33.8738* | No |
| | ASY vs TA | *-9.100191 to 44.87685* | No |
| **Resting BP** | ATA vs NAP | *-5.94766 to 4.502415* | No |
| | ATA vs ASY | *-8.301837 to 0.6393178* | No |
| | ATA vs TA | *-15.45714 to 1.22832* | No |
| | NAP vs ASY | *-7.550255 to 1.330316* | No |
| | NAP vs TA | *-14.71879 to 1.93282* | No |
| | ASY vs TA | *-11.154835 to 4.588982* | No |
| **MaxHR** | ATA vs NAP | *-2.670735 to 11.169202* | No |
| | ATA vs ASY | *14.40914 to 26.24055* | Yes |
| | ATA vs TA | *-9.158105 to 12.929483* | No |
| | NAP vs ASY | *10.19626 to 21.95496* | Yes |
| | NAP vs TA | *-13.387906 to 8.660817* | No |
| | ASY vs TA | *-28.862504 to -8.015809* | Yes |

**Abbreviations**: **ATA:** Atypical Angina , **NAP**: Non-Anginal Pain , **ASY**:Asymptomatic , **TA**: Typical Angina

## AO5:

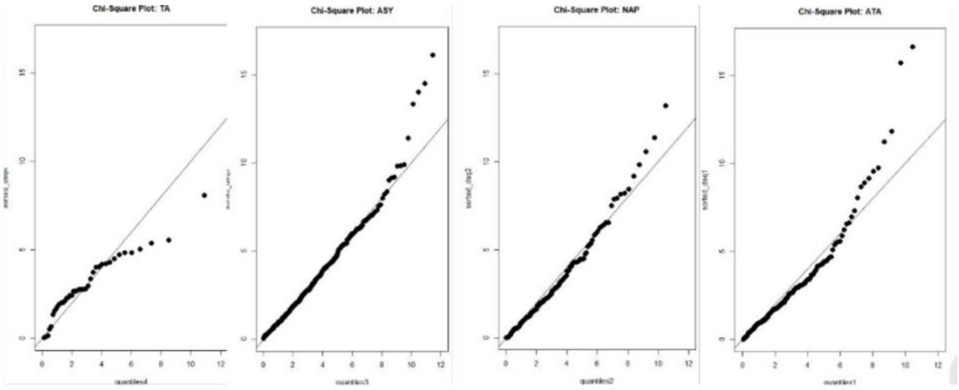


**Image AO5.** Chi-Square Quantile Plots for Different Chest Pain Types

## AO6:

**Table AO6.** One-Way MANOVA, Multivariate Shapiro Wilk test.

| Group | Test Statistic | P-value | Conclusion |
|---|---|---|---|
| (ATA) | *0.96521* | *< 0.001* | Reject Normality |
| (NAP) | *0.94589* | *< 0.001* | Reject Normality |
| (ASY) | *0.97257* | *< 0.001* | Reject Normality |
| (TA) | *0.97405* | *0.5538* | Fail to Reject Normality |

## AO7:

**Table AO7.** One-way ANOVA for each response variable

| Response Variable | Degrees of Freedom (DF) | Sum of Squares (SS) | Mean Square (MS) | F-Value | p-Value |
|---|---|---|---|---|---|
| RestingBP | Between Groups: *3* | *3101* | *1033.8* | *3.496* | *0.0153** |
| | Residuals: *742* | *219425* | *295.7* | | |
| Cholesterol | Between Groups: *3* | *27661* | *9220* | *2.653* | *0.0477** |
| | Residuals: *742* | *2579198* | *3476* | | |
| MaxHR | Between Groups: *3* | *63350* | *21117* | *40.73* | *<0.0001**** |
| | Residuals: *742* | *384717* | *518* | | |
